# Tricuspid regurgitation predicts mortality after liver transplantation in patients with high MELD score: a retrospective cohort study

**DOI:** 10.64898/2026.05.17.26353412

**Authors:** Benjamin C Cailes, Eva-Louise Huber, Claudia R Brick, Avik S Majumdar, Adam G Testro, Marie J Sinclair, Ali Al-Fiadh, James D Theuerle, Julian K Yeoh, Matias B Yudi, Laurence Weinberg, Terase F Lancefield, Anoop N Koshy, Omar Farouque

## Abstract

Tricuspid regurgitation and pulmonary artery systolic pressure may contribute to post-operative morbidity and mortality in liver transplantation. Previous studies suggest that a high Model for End-Stage Liver Disease score may influence the relationship between tricuspid regurgitation and post-operative mortality. Adult patients undergoing liver transplantation workup between 2010 and 2023 were included in this retrospective observational cohort study. Patients with significant portopulmonary hypertension were excluded. Transthoracic echocardiograms were completed pre-transplant and patients were followed up for one year post-operatively. 1031 patients (median MELD score 17, IQR 12-23) underwent transthoracic echocardiography for liver transplantation workup, of whom 708 underwent successful transplantation. Mild or greater tricuspid regurgitation did not predict 1-year mortality in the overall population (HR 1.79 (95% CI 0.78-4.11), p=0.19). Among patients with MELD scores ≥20, mild or greater tricuspid regurgitation was a significant predictor of 1-year mortality (7 (12.7%) vs 9 (3.8%), p=0.01) (HR 3.46 (1.30-10.32), p=0.02). Tricuspid regurgitation in patients with high MELD scores was associated with a trend towards an increased risk of 30-day major adverse cardiovascular events (9 (16.4)% vs 46 (8.1%), p=0.06), driven predominantly by rates of post-operative heart failure (12.7% vs 3.8%, HR 3.66 (95%CI 1.30-10.32), p=0.01). Elevated pulmonary artery systolic pressure was associated with prolonged hospital stay (30 days (14-46) vs 15 days (11-29), p=0.01). Our study confirms that mild or greater tricuspid regurgitation is a significant predictor of 1-year mortality in patients with high MELD scores undergoing liver transplantation. Tricuspid regurgitation severity should be considered during pre-liver transplantation risk stratification.

## 1. Introduction

Liver transplantation (LT) is a lifesaving procedure for patients with end-stage liver disease. However, it remains a high-risk surgery in which post-operative cardiovascular complications are the leading cause of early post-operative morbidity and mortality, as well as non-graft related mortality in the longer term.(1-3) Cardiac risk assessment in LT candidates varies among centers and involves evaluating multiple aspects of cardiac function, including coronary artery disease (CAD), left and right ventricular (RV) function, and valvular lesions.(4-9) Although the importance of CAD and left ventricular systolic function has been studied extensively, limited data exist regarding the prognostic implications of valvular disease in the LT setting.(6) In particular, tricuspid regurgitation (TR) is a lesion of interest because elevated right atrial pressures may exacerbate hepatic congestion or worsen pre-existing liver dysfunction.

Several studies have investigated the effects of pre-existing TR on post-LT outcomes, yielding inconsistent results.(10-12) Two reports have suggested that mild or greater (≥mild) TR is an independent predictor of mortality,(10,11) whereas another study found no association between TR and either 1- or 3-year mortality.(12) A recent meta-analysis identified TR as an independent predictor of post-LT all-cause mortality, but acknowledged that data on this topic are lacking.(13) A high MELD score has previously been hypothesized to influence the hemodynamic significance of tricuspid regurgitant lesions. It is well known that patients with high MELD scores have a more hyperdynamic circulation and increased fluid retention, leading to elevated systemic venous pressures, and a susceptibility to liver injury due to backflow of pressure (14). In the context of hemodynamic stress, there is increased backflow through the tricuspid valve and thus increased backflow into the liver.

Separately, measurement of TR jet velocity acts as an estimate for the pulmonary artery systolic pressure (PASP) using the Bernoulli equation.(15) Significant portopulmonary hypertension (PPH), defined as a form of group 1 pulmonary hypertension associated with cirrhosis and portal hypertension, is known to carry a high operative risk and has poor post-LT outcomes.(16-18) However, whether mild elevations in the PASP (not meeting diagnostic criteria for PPH) also increase perioperative risk is less clear.(10, 11) In this study, we aimed to determine whether the presence of ≥mild TR differentially impacts 1-year mortality among LT recipients with high (≥20) vs low (<20) MELD scores. We further investigated whether non-PPH related elevations in pulmonary pressures (i.e. mild PASP elevations not meeting criteria for PPH) correlate with increased morbidity or mortality.

## 2. Patients and Methods

### 2.1 Patient Population

Consecutive adult patients (≥18 years of age) who successfully underwent LT between 2010 and 2023 at the Victorian Liver Transplant Unit in Melbourne, Australia, were included in this observational cohort study. All demographic and clinical data were entered prospectively at the time of LT assessment and subsequently extracted for the purpose of the current study. In addition, medical history, etiology of liver disease, and severity of liver disease (as assessed by MELD and Child–Pugh scores), were obtained from a prospectively maintained institutional LT database. Diagnosis of cirrhosis was based on histology or on clinical, laboratory and radiological findings. Blood tests as part of the liver transplant assessment included liver function tests, urea, serum electrolytes, and creatinine. Patients who were not transplanted and those undergoing re-transplantation or multi-organ transplantation were excluded, as were those with a diagnosis of PPH. All patients received whole grafts from deceased donors. All patients were abstinent from alcohol for six months before the study. This project received approval from the Austin Health Human Research Ethics Committee (LNR/18/Austin/177).

### 2.2 Imaging Analysis

Echocardiographic images were acquired using a commercially available system and reviewed by an imaging cardiologist as per our institutional protocol. Targeted resting echocardiographic images, including valvular assessment, were obtained according to American Society of Echocardiography guidelines.(15) This included, standard resting echocardiographic images of the left ventricle including parasternal long axis, short axis and apical 4, 3, and 2 chamber views to assess for regional wall motion abnormalities. Right ventricular function was assessed in detail including measurements of systolic function such as the tricuspid annular plane systolic excursion, and RV systolic excursion velocity. Color and continuous-wave Doppler examination of the tricuspid valve, from multiple windows, and Doppler interrogation of the hepatic veins was performed for quantification of TR severity and estimation of PASP. TR severity was graded as trivial, mild, moderate, or severe by the reporting cardiologist. The high reproducibility of echocardiographic parameters in our laboratory has been previously reported.(19, 20)

### 2.3 Outcome Assessment

The primary endpoint for this study was all-cause mortality at one year. A MELD cut-off of 20 was chosen to differentiate high- and low-MELD patients, consistent with previous studies.(10-12) Secondary endpoints included long term mortality to maximal follow-up, the composite outcome of major adverse cardiovascular events (MACE) at 30 days post-LT, length of intensive care unit stay (hours) and hospital stay (days) as well as rates of readmission within 90 days. A 30-day time-period was chosen for quantifying MACE outcomes in accordance with current reporting standards of perioperative outcomes in non-cardiac surgery.(21) MACE was defined as any recorded episodes of acute coronary syndrome, decompensated heart failure or ventricular arrhythmia. Chronic kidney disease is defined as being Grade III chronic kidney disease or lower as defined by the Kidney Disease: Improving Global Outcomes 2024 guidelines, and no patients were on dialysis at the time of transplantation.(22) Primary and secondary endpoints were obtained during long-term follow-up from a prospectively maintained institutional liver transplant database and supplemented by retrospective electronic medical record review by two cardiologists (B.C. and A.N.K.). This followup was conducted until the 31^st^ of December 2024.

### 2.4 Statistical Analysis

Results are expressed as mean ± standard deviation or median with interquartile range if the data were non-normally distributed. This was assessed using skewness and kurtosis tests of normality. Comparisons between groups were performed with the chi-square test for categorical data and the Student t-test or Mann–Whitney U test as appropriate for continuous data. Correlation was assessed using Spearman’s rank correlation. Kaplan-Meier curves were produced using survival-time analyses and hazard ratios (HR) calculated using a Cox proportional-hazards regression model. All reported P values are 2-tailed, with values <0.05 considered significant.

Statistical analyses were performed using Stata 17/MP (StataCorp, College Station, TX).

## 3. Results

### 3.1. Baseline Characteristics

A total of 1031 patients underwent pre-operative transthoracic echocardiography (TTE) as part of LT assessment, of whom 708 (68.7%) proceeded to successful liver transplantation (Figure 1). The median time from TTE to transplantation was 174 days (IQR 82-356 days). All patients underwent tricuspid valve assessment during the TTE, with 118 patients (16.7%) diagnosed as having ≥mild TR (mild = 100 (84.7%), moderate = 11 (9.3%), severe = 7 (5.9%)). The baseline characteristics of patients with and without ≥mild TR are presented in Table 1. Patients with ≥mild TR were largely similar to those with no or trivial TR with regards to age, sex and cardiovascular risk factors. A total of 49 patients had HCC recorded as their primary indication for transplantation. Of these, 13 (26.5%) had mild or greater TR. The median MELD score in the study population was 17, however, patients with ≥mild TR had a significantly higher median MELD score than those without (20 (15-26) vs 17 (12-22), p=0.01), and were also more likely to have a history of atrial fibrillation (9.3% vs 3.4%, p=0.01). When divided into high and low MELD groups, 290 patients (41.0%) had a recorded MELD score of ≥20 (55 (19.0%) with ≥mild TR and 235 (81.0%) with <mild TR), and 418 (59.0%) had a score <20 (63 (15.1%) with ≥mild TR and 355 (84.9%) with <mild TR).

**Table 1.** Baseline characteristics in patients with and without tricuspid regurgitation.

|  | ≥Mild TR (n=118) | <Mild TR (n=590) | p-value |
| --- | --- | --- | --- |
| Age (Years) | 56.7 (51.9-62.0) | 58.0 (49.7-63.4) | 0.69 |
| Sex (Male) | 74 (62.7%) | 407 (69.1%) | 0.19 |
| Liver Pathology |  |  |  |
| HBV or HCV | 34 (28.8%) | 195 (33.1%) | 0.37 |
| MASH | 11 (9.3%) | 75 (12.7%) | 0.30 |
| Alcohol related liver disease | 21 (17.8%) | 116 (19.7%) | 0.64 |
| HCC | 13 (11.0%) | 36 (6.1%) | 0.06 |
| MELD Score | 20 (15-26) | 17 (12-22) | **0.01** |
| Dialysis | 0 | 0 |  |
| Creatinine | 1.02 (0.76-1.35) | 0.94 (0.76-1.28) | 0.51 |
| International Normalized Ratio | 1.6 (1.3-2.0) | 1.4 (1.2-1.8) | 0.63 |
| Bilirubin | 5.67 (3.04-15.09) | 3.16 (1.35-7.25) | **<0.001** |
| Na | 136 (131-139) | 137 (134-140) | 0.24 |
| CAD | 7 (5.9%) | 24 (4.1%) | 0.37 |
| CCF | 3 (2.5%) | 6 (1.0%) | 0.18 |
| Hypertension | 22 (18.6%) | 119 (20.2%) | 0.15 |
| Dyslipidaemia | 29 (24.6%) | 104 (17.6%) | 0.09 |
| Diabetes | 40 (33.9%) | 188 (31.9%) | 0.67 |
| Insulin | 17 (14.1%) | 71 (12.0%) | 0.48 |
| Smoking History | 62 (54.5%) | 319 (54.1%) | 0.76 |
| Atrial Fibrillation | 11 (9.3%) | 20 (3.4%) | **0.01** |
| Chronic Kidney Disease | 2 (1.7%) | 28 (4.8%) | 0.13 |

CAD; Coronary artery disease, CCF; Congestive cardiac failure, TR; Tricuspid regurgitation.

**Figure 1.**
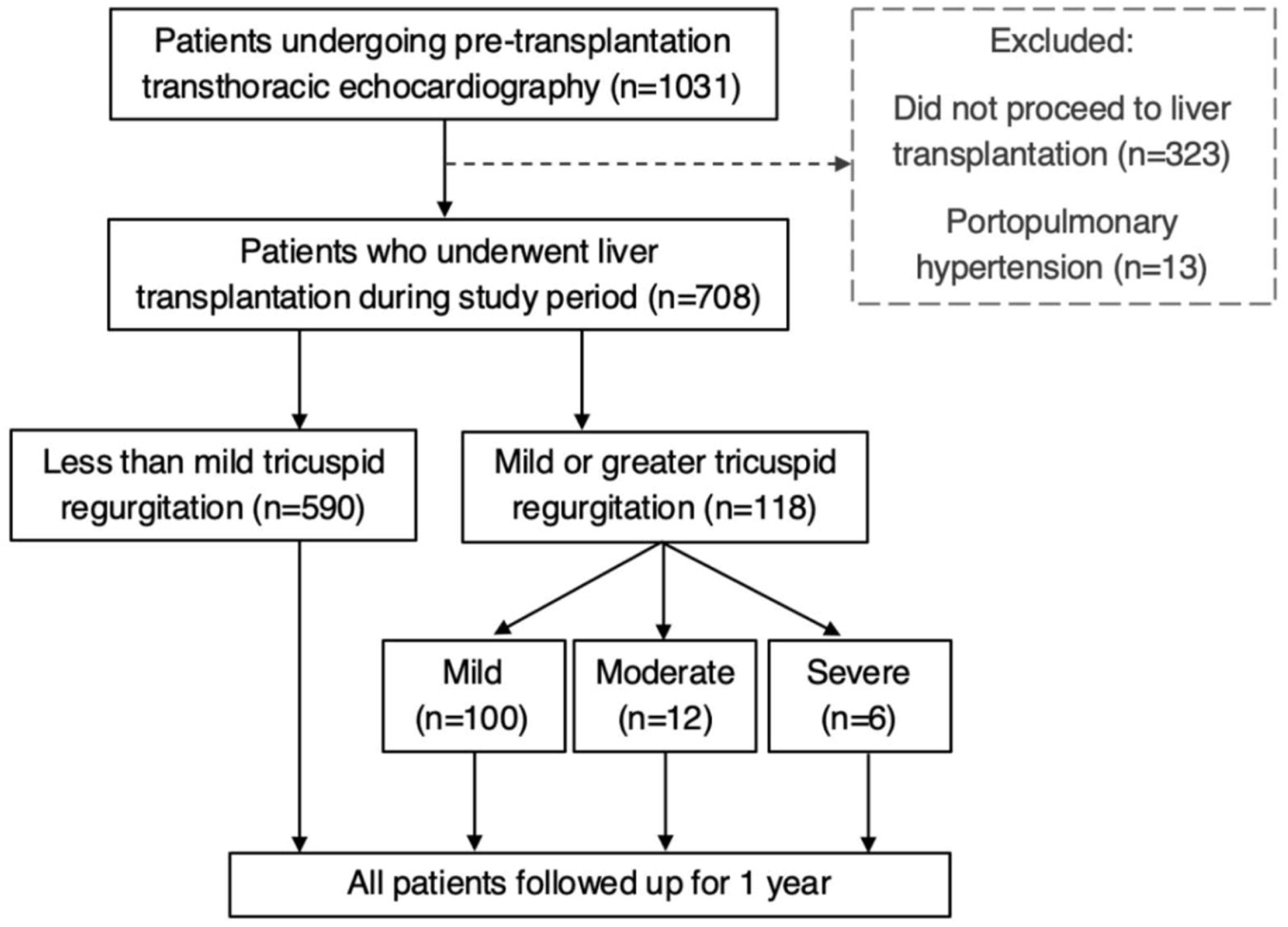
Study details and patient inclusion.

### 3.2. Cardiac Outcomes

#### 3.2.1. Mortality

Predictors of one-year mortality are described in Table 2. Thirty-one patients (4.4%) died during follow-up with 677 (95.6%) surviving. In the overall study population, the presence of ≥mild TR was not significantly associated with 1-year mortality, with 8 deaths (6.8%) seen among those with ≥mild TR and 23 deaths (3.9%) seen among those with <mild TR (p=0.16). However, in patients with MELD scores ≥20, ≥mild TR was associated with a significantly increased risk of 1-year mortality (7 (12.7%) vs 9 (3.8%), p=0.01) (HR 3.66 (1.30-10.32), p=0.02) (Table 2) (Figure 2). Similarly, these high-MELD patients had a significantly increased risk of long-term mortality to maximal follow-up (15 (27.3%) vs. 30 (12.8%), p=0.01) (HR 2.56 (1.26-5.19), p=0.01).

**Table 2.** Predictors of one-year mortality.

|  | **Died (n=31)** | **Survived (n=677)** | **HR (95% CI)** | **p-value** |
| --- | --- | --- | --- | --- |
| Age (Years) | 59.7 (56.5-62.5) | 57.5 (50.3-63.2) | 0.001 (-0.001-0.003) | 0.19 |
| Sex (Male) | 22 (71.0%) | 459 (67.8%) | 1.16 (0.53-2.56) | 0.14 |
| MASH | 2 (6.5%) | 84 (12.4%) | 0.49 (0.11-2.08) | 0.28 |
| Hepatitis B/C | 6 (19.4%) | 223 (32.9%) | 0.49 (0.20-0.21) | 0.10 |
| MELD | 19 (14-23) | 17 (12-23) | 0.002 (-0.001-0.003) | 0.10 |
| CAD | 2 (6.5%) | 29 (4.3%) | 1.54 (0.35-6.77) | 0.58 |
| Congestive cardiac failure | 1 (11.1%) | 29 (4.2%) | 2.87 (0.35-23.76) | 0.39 |
| Hypertension | 4 (12.9%) | 137 (20.2%) | 0.58 (0.20-1.70) | 0.29 |
| Dyslipidemia | 8 (25.8%) | 125 (18.5%) | 1.541 (0.67-3.51) | 0.33 |
| Diabetes | 8 (25.8%) | 220 (32.5%) | 0.72 (0.32-1.64) | 0.43 |
| Insulin | 4 (12.9%) | 84 (12.4%) | 1.05 (0.36-3.06) | 0.94 |
| Smoking | 13 (41.9%) | 368 (54.4%) | 1.61 (0.29-1.26) | 0.18 |
| Atrial fibrillation | 2 (6.5%) | 29 (4.3%) | 1.54 (0.35-6.77) | 0.59 |
| Chronic kidney disease | 2 (6.5%) | 28 (4.1%) | 1.60 (0.36-7.01) | 0.56 |
| TR ≥ mild | 8 (6.8%) | 110 (93.2%) | 1.79 (0.78-4.11) | 0.19 |
| MELD <20 | 1 (1.6%) | 62 (98.4%) | 0.39 (0.05-3.04) | 0.31 |
| MELD ≥20 | 7 (12.7%) | 48 (87.3%) | 3.66 (1.30-10.32) | **0.02** |
| TR velocity >2.8m/s | 8 (28.6%) | 123 (23.1%) | 1.33 (0.57-3.09) | 0.52 |
| Mitral regurgitation ≥ mild | 5 (16.1%) | 85 (12.5%) | 1.34 (0.50-3.58) | 0.57 |
| Aortic stenosis ≥ mild | 2 (6.5%) | 17 (2.5%) | 2.68 (0.59-12.14) | 0.25 |
| Tricuspid annular plane systolic excursion | 23.5 (18-28) | 26 (21-30) | 0.93 (0.85-1.01) | 0.10 |
| RV systolic excursion velocity | 14 (12-18) | 15 (13-18) | 1.01 (0.88-1.16) | 0.89 |

CAD; Coronary artery disease, TR; Tricuspid regurgitation.

**Figure 2.**
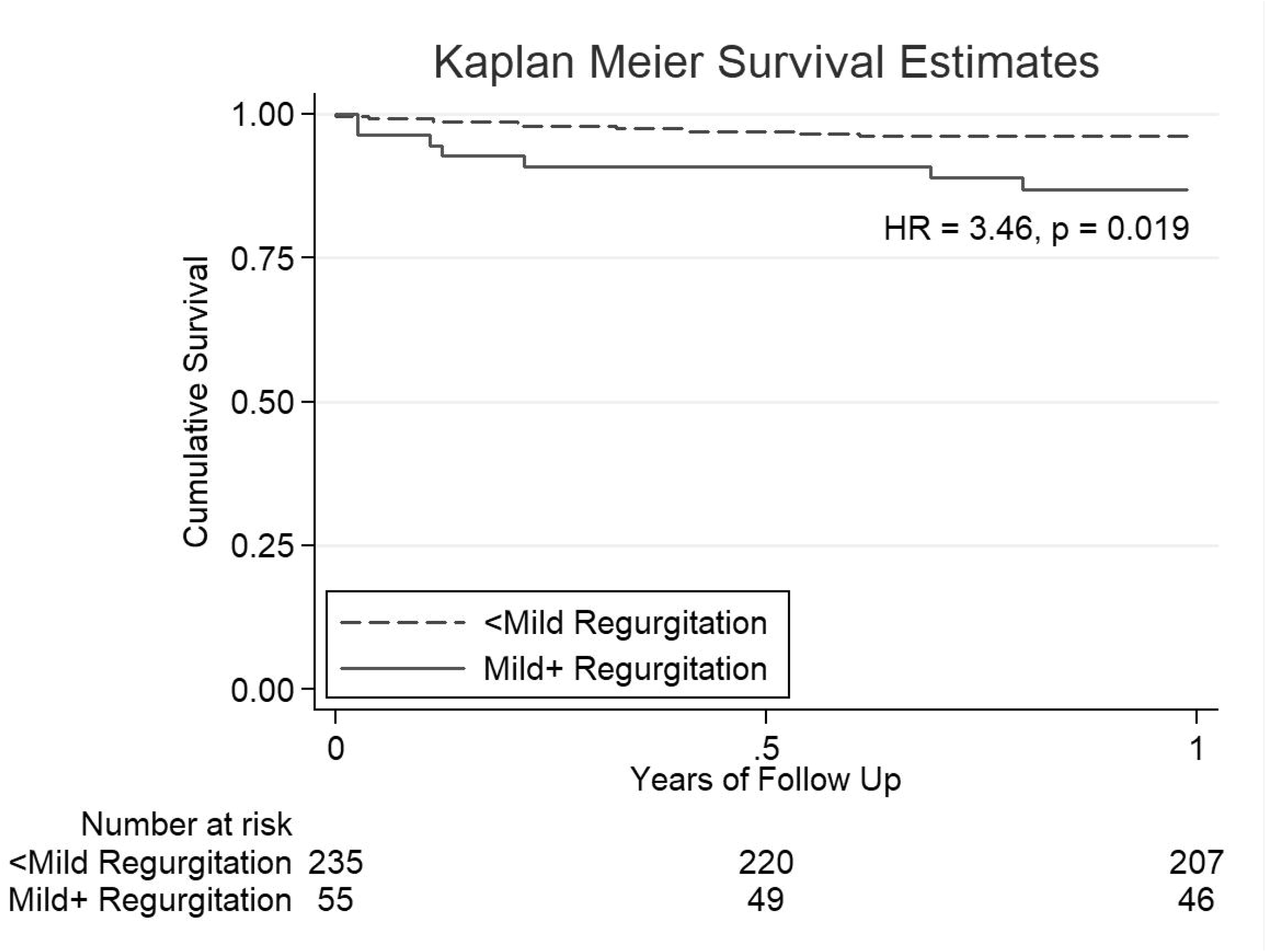
Kaplan-Meier curves demonstrating survival post liver transplant in tricuspid regurgitation with MELD ≥20 at 1 year.

A sub-analysis was performed looking exclusively at mild tricuspid regurgitation, and excluding patients with moderate or severe TR. Among patients with a MELD≥20 and mild TR, 7 out of 47 patients (14.9%) died within one-year of LT, compared to only 9 of 243 patients with no TR (3.7%, p=0.01) (HR 4.39 (95%CI 1.55-12.47), p=0.01). In contrast, patients with MELD <20 did not demonstrate an association with mortality (1.9% vs. 3.9%, p=0.46) (HR 0.47 (95%CI 0.06-3.64), p=0.42). Among patients with HCC recorded as their primary indication for transplant, and who had mild or greater TR, none died within one year of LT (0.0%), while three mortality events were seen among patients with HCC but no TR (8.3%), p=0.28. When patients with HCC were excluded, patients with ≥mild TR still demonstrated a trend towards an increased risk of one year mortality (8 (7.6%), vs. 20 (3.6%), p=0.62), (HR 2.20 (0.94-5.14), p=0.09). This was once again driven by a significant association between TR and mortality among patients with a high MELD score ≥20 (7 (12.7%) vs 9 (3.9%), p=0.01) (HR 3.60 (1.28-10.13), p=0.02). Among patients with specifically mild TR (excluding those with moderate or severe TR as well as those with HCC), a significant difference in rates of one year mortality was seen for the population as a whole, (3 (17.7%) vs 3 (4.1%), p=0.04) (HR 2.57 (1.10-6.03), p=0.04). This was once again driven by a strong association among patients with MELD ≥20 (7 (14.9%) vs 9 (3.8%), p=0.01) (HR 4.47 (1.58-12.69), p=0.01), with no association seen among patients with MELD <20 in isolation (p=0.69).

#### 3.2.2. MACE, Post-Operative Hospital Stay, Readmission, and Graft Outcomes

A total of 52 MACE events occurred within 30-days of LT, affecting 7.3% of the cohort. In the overall cohort, ≥mild TR did not predict 30-day MACE (12 (10.2%) vs 40 (6.8%), p=0.20). However, among high MELD patients with ≥mild TR, a trend towards an increased risk of MACE was observed (9 (16.4%) vs 46 (8.1%), p=0.06), primarily driven by significantly higher rates of post-operative heart failure (12.7% vs 3.8%, HR 3.66 (95%CI 1.30-10.32), p=0.01), as seen in Table 3. No significant differences were observed in rates of cardiac arrhythmias or acute coronary syndromes. Additionally, in high MELD patients (≥20), 90-day readmission rates were significantly higher than in patients with no or trivial TR (13 (23.6%) vs 28 (11.9%), p=0.03, HR 2.29 (95% CI 1.10-4.78), p=0.03), and a trend was seen towards prolonged length of hospital stay (19 days (13-39) vs 15 days (11-28), p=0.09).

**Table 3.** Major adverse cardiovascular events amongst patients with mild or greater tricuspid regurgitation.

|  | ≥Mild TR | <Mild TR | p-value |
| --- | --- | --- | --- |
| Patients with MELD ≥20: |  |  |  |
| MACE | 9 (16.4%) | 19 (8.1%) | 0.06 |
| Heart failure | 7 (12.7%) | 9 (3.8%) | **0.01** |
| Acute coronary syndrome | 1 (1.8%) | 4 (1.7%) | 0.95 |
| Ventricular arrhythmia | 1 (1.8%) | 8 (3.4%) | 0.54 |
| Patients with MELD <20: |  |  |  |
| MACE | 3 (4.8%) | 21 (5.9%) | 0.72 |
| Heart failure | 1 (1.6%) | 6 (1.7%) | 0.95 |
| Acute coronary syndrome | 1 (1.6%) | 5 (1.4%) | 0.91 |
| Ventricular arrhythmia | 1 (1.6%) | 13 (3.7%) | 0.40 |

MACE; Major adverse cardiovascular events, TR; Tricuspid regurgitation.

A non-significant trend towards an increased risk of MACE was also seen among patients with a TR velocity of >2.8m/s, suggestive of an elevated PASP (15 (11.5%) vs 28 (6.5%), p=0.06), with the caveat that pulmonary pressures were only able to be estimated in 560 of the 708 patients due to insufficient TR in the remainder. After adjusting for covariates predicting MACE, TR velocity >2.8m/s was not an independent predictor of cardiac outcomes (p=0.17). However, these patients experienced significantly longer overall hospital stays (30 days (14-46) vs 15 days (11-29), p=0.01), as well as higher rates of post-operative acute kidney injury (102 (77.9%) vs 265 (61.8%), p=0.01). No difference was seen in rates of 90-day readmissions (22 (16.8%) vs 64 (14.9%), p=0.60). No significant difference was noted in rates of rejection between patients with ≥mild TR and those without TR (20 (29.4%), vs 36 (21.4%), p=0.19). Similarly, no difference was noted in the rates of biliary strictures occurring within 180 days (16 (23.5%) vs 36 (21.4%), p=0.72).

#### 3.2.3 Severe TR outcomes

While the current study was not powered to explore an association between severe TR and post-operative events, exploratory data regarding the six patients in our cohort with severe TR on pre-operative TTE are listed in Table 4. It was noted that, of the four of these patients who had a repeat TTE prior to transplant, three out of four had reductions in degree of TR with diuresis alone. A minority of patients had a post-operative TTE, thus few conclusions can be drawn regarding the direct effect of liver transplantation on TR.

**Table 4.**
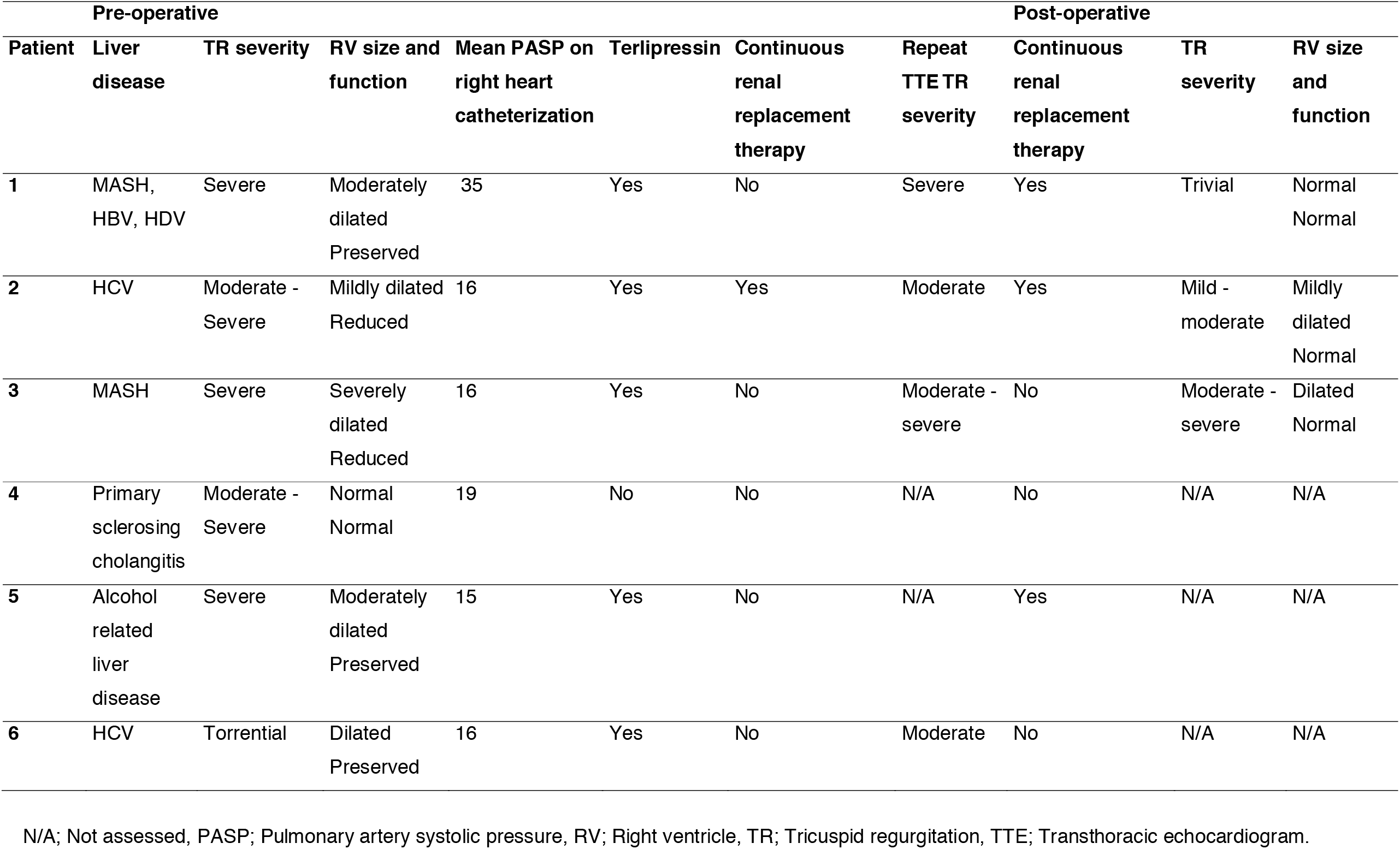
Exploratory data on patients with severe tricuspid regurgitation.

## 4. Discussion

This study represents the largest cohort to date investigating the impact of tricuspid regurgitation in liver transplantation. Our findings confirm that ≥mild TR is independently associated with increased 1-year mortality and post-operative heart failure exacerbations among patients with a high MELD score ≥20. This supports prior hypotheses that TR has greater prognostic relevance in this subset of LT recipients. Following multivariate analysis, controlling for factors including liver disease severity and aetiology, TR was demonstrated to be the strongest predictor of one-year mortality in high MELD patients. Additionally, we observed an association between elevated pulmonary pressures (TR velocity >2.8m/s) and prolonged hospital stay, though no significant relationship was found with 30-day MACE, intensive care unit stay, or 90-day readmission rates (Figure 3).

**Figure 3.**
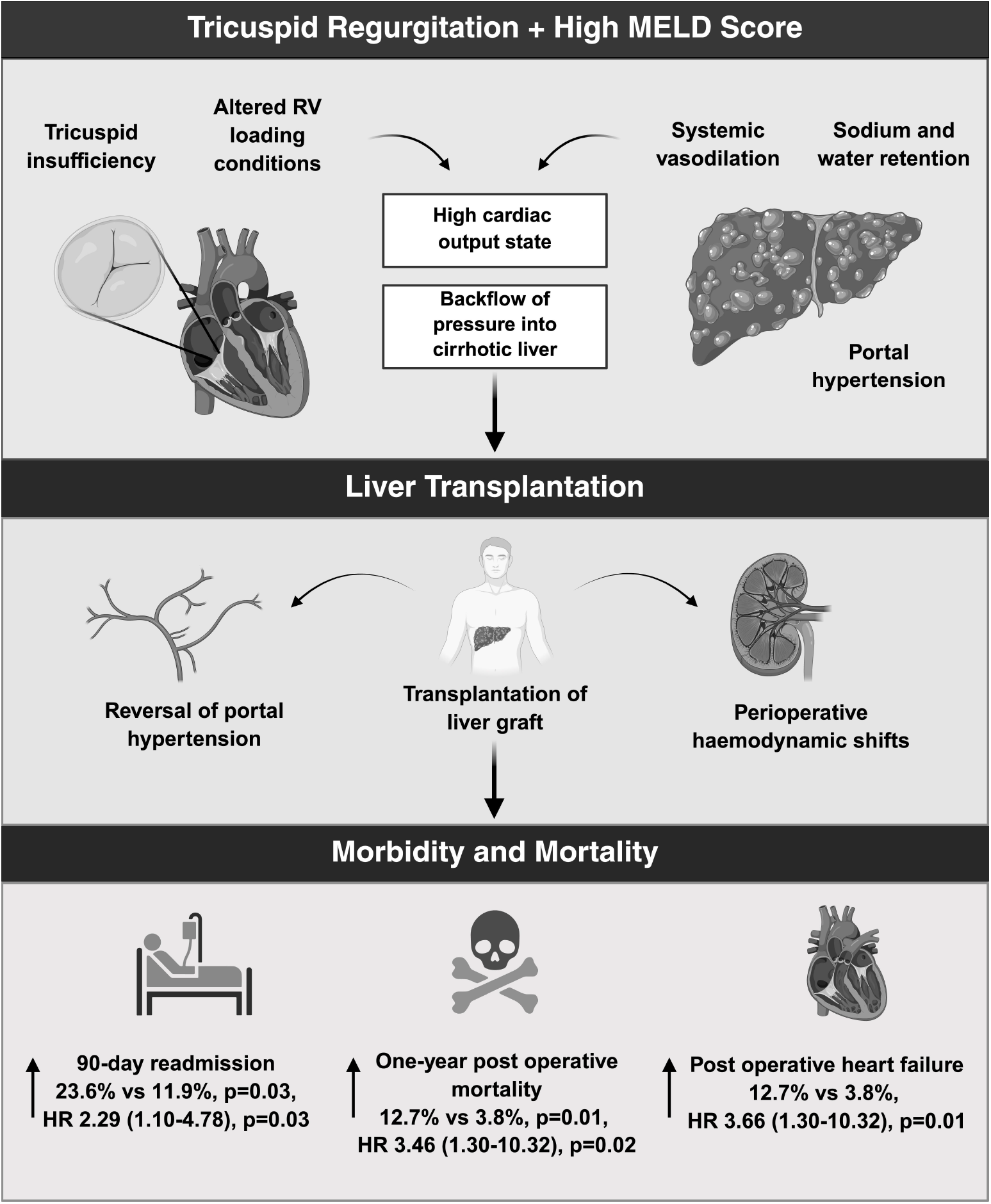
Proposed mechanisms of the association between tricuspid regurgitation and Model for End-Stage Liver Disease with elevated one-year post-operative mortality, heart failure, and hospital readmission. RV; Right ventricle.

In contrast to prior studies by Kia et al and Bushyhead et al, we did not identify an association between TR and mortality in the overall cohort, with our findings being more in line with those of Leithead et al. It has been hypothesized following these prior studies that the variation in median population MELD scores for these may have contributed to the inconsistencies in these results.(12) It is therefore of significance that, when focusing on patients with a MELD of ≥20, those with ≥mild TR demonstrated a significantly increased risk of 1-year mortality, independent of other risk factors. This suggests that TR may contribute to adverse outcomes predominantly in patients with advanced liver disease, potentially due to their more hyperdynamic circulation and hence a greater susceptibility to perioperative hemodynamic stress.

The association between TR and mortality in high MELD patients is biologically plausible. High MELD patients exhibit greater circulatory derangements, including systemic vasodilation, high cardiac output states, and altered RV loading conditions. In this setting, TR may worsen hepatic congestion, impair cardiac reserve, and exacerbate post-operative right heart failure, a pattern reflected in our findings.

Moreover, severity of TR in LT patients may be more dynamic than previously appreciated. Even mild TR, usually considered benign, could be significantly exacerbated by massive intraoperative hemodynamic shifts and fluid resuscitation leading to acute RV overload, compromised systemic perfusion and graft congestion. This dynamic progression could partially explain why high MELD patients with TR had worse outcomes, particularly in the early post-operative period. Moderate and severe TR have been found to independently predict mortality in patients with pulmonary hypertension, supporting the notion of TR being associated with mortality in patients with altered hemodynamic states (23, 24).

Our results help to clarify the relationship between TR and mortality, and confirm that this relationship is modulated by severity of liver disease. This is consistent with prior studies, where the two prior studies that reported an association between TR and increased mortality had high median MELD scores (22.4 and 25, respectively)(10, 11), while conversely, a third study with a lower median MELD of 15 did not find a mortality association.(12) This pattern again suggests that the prognostic impact of TR is more pronounced in patients with more advanced liver disease, reinforcing the importance of MELD-stratified risk assessment.

Unlike previous reports, we did not find an association between PASP elevation and increased MACE risk. This discrepancy may be explained by post-LT reductions in PASP, a well-documented phenomenon that occurs as portal hypertension resolves following transplantation, reducing loading of the RV. Additionally, our cohort had a lower median MELD score (17) compared to prior studies, which may have attenuated the relationship between pulmonary pressures and post-transplant cardiovascular complications. However, our finding that elevated PASP correlated with longer hospital stays supports the notion that these patients remain at heightened perioperative risk among patients who did not meet formal diagnostic criteria for portopulmonary hypertension.

### 4.1. Clinical Implications

Our findings highlight the need to consider the prognostic implications of tricuspid regurgitation in LT risk stratification, particularly in high MELD patients. While traditionally viewed as incidental, even mild TR may become clinically significant under the extreme hemodynamic shifts of LT, contributing to RV volume overload, systemic and graft hypoperfusion, and adverse clinical outcomes. Additionally, TR could potentially mask subclinical RV dysfunction, making standard echocardiographic assessments less sensitive in detecting early RV impairment. This is particularly relevant in LT candidates, where RV function plays a critical role in managing post-operative circulatory changes. Given its association with increased mortality and post-operative complications, TR should not be overlooked during pre-transplant assessment. Additionally, the prolonged hospital stays seen in patients with elevated PASP further reinforce the need for close perioperative monitoring of this sub-group of patients. Future studies should explore whether optimizing right heart function through volume management, pulmonary vasodilators, or pre-LT transcatheter interventions, could improve post-transplant outcomes in high-risk patients.

The strengths of this study include the large cohort size, making it the most comprehensive analysis of TR in LT recipients to date, with rigorous echocardiographic assessment and independent adjudication of MACE outcomes enhancing reliability. However, certain limitations warrant discussion. Firstly, as a retrospective single center study, it is subject to selection and reporting biases. However, two independent reviewers undertook an individual case review to ensure accurate identification of MACE outcomes. Secondly, TR severity was assessed at a single preoperative time point, which may not reflect the dynamic changes that occur during LT, particularly given the large fluid shifts and hemodynamic stress of transplantation. However, this is the timepoint at which risk stratification occurs and hence we believe is the most clinically relevant timepoint to assess. Finally, our study was not powered to evaluate the impact of severe TR on outcomes or the ability of potential interventions to mitigate this risk. Despite these limitations, our findings underscore the need for heightened awareness of TR in high MELD patients and support further investigation into targeted perioperative management strategies.

## 5. Conclusion

This study confirms, for the first time, the hypothesis that ≥mild TR is a significant predictor of 1-year mortality for high MELD patient populations undergoing liver transplantation. These findings underscore the need to consider TR severity during LT risk stratification and highlight the potential value of interventions aimed at optimizing right heart function preoperatively. Further research is warranted to explore whether targeted management of TR could improve post-transplant outcomes in high-risk patients.

## Data Availability

All data produced in the present study are available upon reasonable request to the authors.

## Abbreviations

CAD: Coronary Artery Disease
LT: Liver Transplantation
MACE: Major Adverse Cardiovascular Events
N/A: Not Assessed
PASP: Pulmonary Artery Systolic Pressure
PPH: Portopulmonary Hypertension
RV: Right Ventricle
TR: Tricuspid Regurgitation
TTE: Transthoracic Echocardiography

## Acknowledgments

Assistance with the study: none.

Presentation: none.

---

Figure 3 created in BioRender. Koshy, A. (2026) https://BioRender.com/zyhem18

