## Supplementary Table 1 for "Tricuspid regurgitation predicts mortality after liver transplantation in patients with high MELD score: a retrospective cohort study"

| **Supplementary Table 1.** Components of the MELD score among patients with mild or greater tricuspid regurgitation. | | | |
| --- | --- | --- | --- |
|  | MELD <20 (n=63) | MELD ≥20 (n=55) | p-value |
| MELD Score  Dialysis  Creatinine  International Normalized Ratio  Bilirubin  Sodium | 15 (13-17)  0  0.85 (0.72-1.13)  1.4 (1.2-1.6)  3.22 (1.7-4.85  138 (131-140) | 27 (22-31)  0  1.16 (0.84-1.55)  2.0 (1.7-2.6)  13.98 (7.08-24.62)  135 (131-137) | **<0.001**  **0.01**  **<0.001**  **<0.001**  0.44 |
